# Physicians’ experiences with telemedicine after the publication of India’s Telemedicine Practice Guidelines

**DOI:** 10.1101/2024.02.10.24302616

**Authors:** Vikranth Harthikote Nagaraja, Biswanath Ghosh Dastidar, Shailesh Suri, Anant R. Jani

**Affiliations:** Department of Engineering Science, Institute of Biomedical Engineering, University of Oxford, Oxford OX1 3PJ, UK; Centre for Human Movement and Rehabilitation, School of Health and Society, University of Salford, Salford, M5 4WT, UK; Oxford India Centre for Sustainable Development, University of Oxford, Oxford OX2 6HD, UK; GD Institute for Fertility Research, Kolkata – 700025, India; Cambridge Reproduction, Department of Physiology, Development and Neuroscience, Downing Street, Cambridge CB2 3EL, UK; Department of Obstetrics and Gynaecology and Centre of Excellence (CoE) in IVF–ART, Institute of Postgraduate Medical Education & Research (IPGMER) and SSKM Hospital, Kolkata – 700020, India; Heidelberg Institute for Global Health, Heidelberg, Germany

**Keywords:** Digital health, Internet-based survey, Healthcare delivery, Qualitative analysis, Healthcare policy, India, Telemedicine

## Abstract

**Background:** Digital health is an important factor in India’s healthcare system. Inclusive policy measures, a fertile technological landscape, and relevant infrastructural development with unprecedented levels of telemedicine adoption, catalysed by the COVID-19 pandemic, have thrown open new possibilities and opportunities for clinicians, end-users, and other stakeholders. Nevertheless, several challenges remain in properly integrating and scaling telemedicine in India. This study’s objective was to understand the views of practising physicians in India on the use of telemedicine, with a particular focus on the period after the release of India’s Telemedicine Practice Guidelines in India, after which telemedicine was rapidly implemented.

**Methods:** We acquired data through an anonymous, internet-based survey (with a cross-sectional time perspective) of 444 physicians across India. These responses were subjected to qualitative data analysis (via inductive coding and thematic analyses) and descriptive statistics, as appropriate.

**Results:** Most responses (n=51) were categorised under a code indicating that telemedicine-led healthcare delivery compromised treatment quality. The second largest proportion of responses (n=22) suggested that ‘Accessibility, quality and maturity of software and hardware infrastructure’ was a considerable challenge.

**Conclusions:** Despite the considerable uptake, perceived benefits, and the foreseen positive role of telemedicine in India in delivering universal coverage, several challenges of telemedicine use (viz., technical, user experience-based integration, and non-user-based integration challenges) have been identified, which need to be addressed to realise telemedicine’s potential. Several relevant opportunities are suggested to inform policy and practice to ensure effective utilisation of telemedicine in India.

## 1. Introduction

Digital determinants of health (DDoH) are an essential component of 21^st^-century healthcare [1,2]. DDoH consist of digital infrastructure as well as telemedicine, mobile phones and apps, wearable devices, sensors and extended reality [3,4]. If used well, DDoH can transform care pathways to increase access and equity while improving health outcomes [3]. India, the world’s most populous country, is amongst the fastest-digitising nations and is making important strides in addressing DDoH [5]. The National Digital India initiative was launched in 2015 with the stated aim “to transform India into a digitally empowered society and knowledge economy” [6]. As of 2021, *Aadhaar*, the world’s largest digital identity program, has around 1.3 billion users [7], and the BharatNet programme aims to extend a fibre network to approximately 250,000 villages by 2025, driving the estimated number of smartphone users to one billion by 2026 [8].

In healthcare, the 2020 National Digital Health Mission (NDHM) Initiative was launched to support the goal of Universal Healthcare through technology [9]. NDHM proposed a national digital architecture of key components, including a national health identifier for all citizens (Ayushman Bharat Health Account [ABHA]), digitised patient health records, and a centralised patient-consent mechanism to allow sharing of health information [10]. The rollout of telehealth in India was boosted by the COVID-19 lockdowns. Besides releasing Telemedicine Practice Guidelines, the free Telemedicine Service, eSanjeevani, was rolled out nationally in 2019 and has since been used by over 148 million patients [11,12]. Despite the impressive progress in digitising healthcare, several challenges impede the more rapid and widespread rollout of digital health. These challenges include the lack of adequate infrastructure, especially in rural areas; inequity of access; the continued commitment of the Indian Government and other institutional stakeholders; patient satisfaction; system usability; and buy-in of frontline clinicians [13–15].

In this manuscript, we present survey findings of physicians across India who were delivering services after the publication of the Telemedicine Practice Guidelines in India. Our aim was to better understand their views on telemedicine use and the challenges they experienced. By understanding these perspectives, we hope the results of our study can inform the future design of effective telemedicine policy and practice with the ultimate goal of realising universal healthcare in India.

## 2. Materials and Methods

### 2.1 Study design

The methods and study findings adhere to the **Che**cklist for **R**eporting Results of **I**nternet **E**-**S**urveys (**CHERRIES**) [16]. Best practices and guidelines on conducting and reporting survey research were also followed [17]. Further details of the study design are as previously reported [18].

### 2.2 Ethical approval

This study surveyed physicians anonymously via an internet-based questionnaire to understand their experiences and perceptions of telemedicine. No confidential or personal information was solicited from survey respondents, and patients were not involved in any way in the study. Given this, and as per the Declaration of Helsinki of the World Medical Association, no ethical approval was necessary. Furthermore, this study was classified as a ‘service review’ when consulted with the institutional ethics committee at the University of Oxford (confirmation available upon request).

### 2.3 Survey design

An open, self-administered, anonymous internet-based survey [18] was used to gather quantitative and qualitative data. Multiple-choice and open-ended questions on telemedicine use were included. Specifically, two free-text questions were incorporated to cross-sectionally understand the perceived challenges with telemedicine use. Apart from questions that needed free-text inputs, all questions were compulsory. Where appropriate, adaptive questioning, response validation, and/or non-response options were used.

### 2.4 Sampling strategies and target population

A non-probability sampling approach was used. Participation was voluntary, and study participants were notified that all data gathered was non-identifiable and would only be used for research purposes. Reimbursements were not provided to survey respondents.

### 2.5 Data collection

*JISC Online Surveys* (https://www.onlinesurveys.ac.uk/), a University of Oxford-approved survey tool, was used to prepare, host, and administer the online survey between 09/12/2020 and 28/02/2021. Responses were recorded on an automated and password-protected database, stored on secure JISC Online Surveys, as well as an institutionally issued personal computer of the study co-author (VHN). The survey responses corresponding to fully completed questionnaires were exported from the aforementioned survey tool in comma-separated values (.CSV) format for offline analysis.

### 2.6 Data analysis

Microsoft Excel 2016 was used for quantitative data analysis. Google Sheets were used to aggregate, pre-process and qualitatively analyse replies to the two optional open-ended questions (on perceived challenges with telemedicine and additional feedback) [19]. Only complete and relevant survey responses were considered.

Pre-processing steps included the following: (i) some open-ended entries had to be split into more than one response as they spanned multiple/unrelated codes, and (ii) some responses (that were incomplete, had a single-word entry, etc.) were removed, leading to 116 eligible responses. Consensus on eligibility was achieved for all open-ended responses by the authors.

Coding and thematic analyses were used for qualitative data analysis [20–22]. A deductive coding approach was first used. This was based on the constructs per a relevant review article [23], with the authors assigning the open-ended survey responses to these codes. After coding, it was found that the authors agreed 59 times and disagreed 57 times. The authors realised that the codes were not adequately suited to the survey responses because (i) the codes/constructs were not appropriate for our study, which was conducted in a low-resource setting; (ii) the considered study [23] is over six years old; and (iii) the rapid of adoption of telemedicine in our study period rendering some of the codes and constructs from Langbecker et al. [23] obsolete.

The authors then adopted an inductive approach because its emergent nature was more appropriate for the unique situation of unprecedented levels of telemedicine use after the release of the Telemedicine Practice Guidelines [11] in India. Codes were identified by utilising (i) *In Vivo* coding, (ii) *Open* coding, and (iii) *Descriptive* coding. The authors read through the survey responses, applied codes independently, and then performed a second iteration of coding that added an interpretive lens. In total, three rounds were necessary, and 11 codes were agreed upon (Figure 1). Any conflict in coding was resolved by consensus [21].

**Fig. 1.**
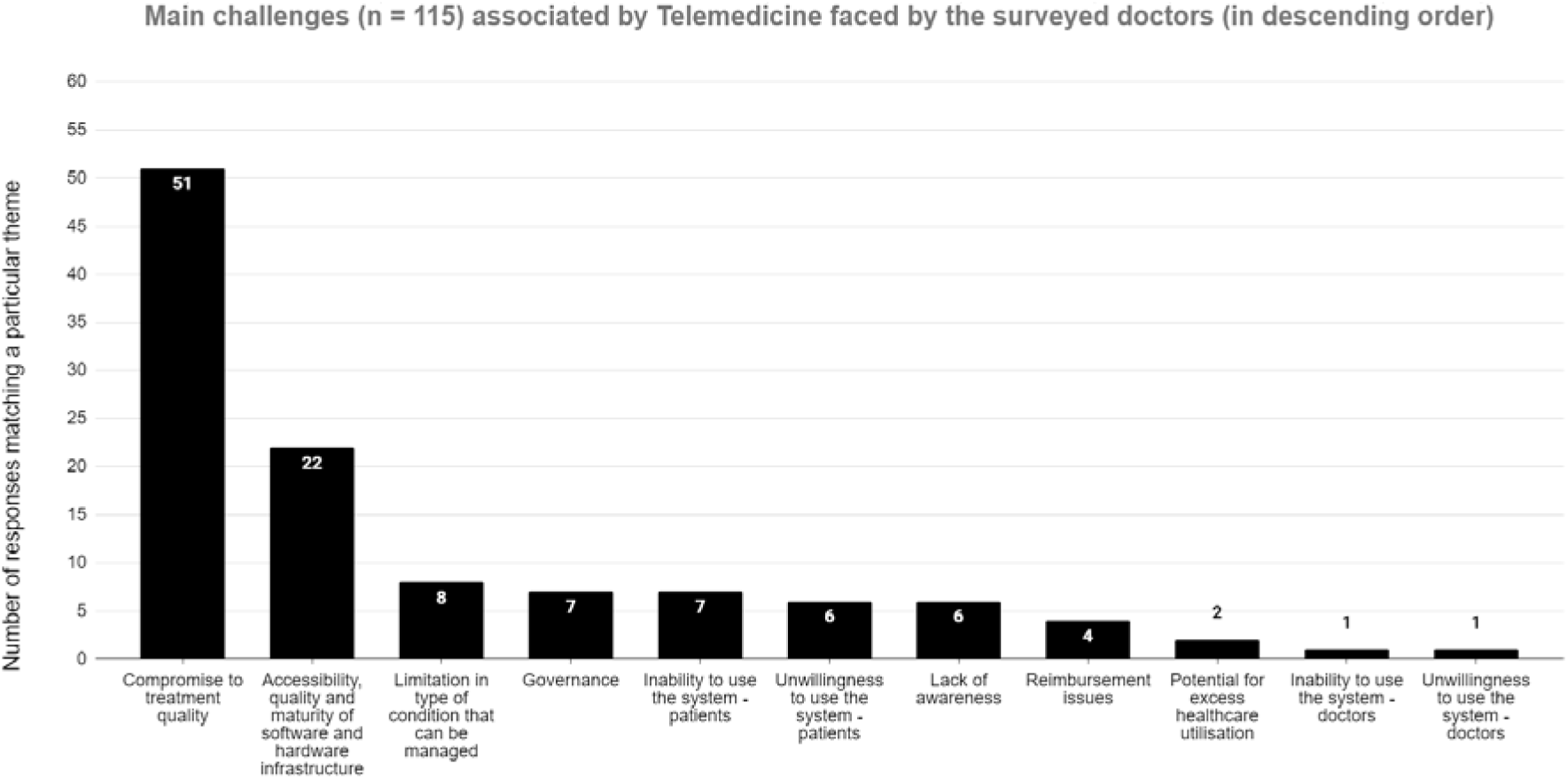
Pareto histogram of the inductive coding summary of challenges associated with telemedicine use faced by physicians in India.

After the first inductive coding round, the authors agreed 89 times and disagreed 27 times. After forming a consensus in the second coding round, all but one response saw agreement (i.e., 115 agreements).

## 3. Results

As reported previously [18], survey respondents generally represented the distribution of physicians across India, with 81 % being from urban areas, 17 % from semi-urban areas, and 2 % from rural areas. 91.9 per cent of survey respondents were actively involved in the frontline around the time of the first wave of the COVID-19 pandemic. Participants were asked about their perceptions of the use and challenges linked to telemedicine in India.

### 3.1 Telemedicine use

For the period after the publication of the Telemedicine Practice Guidelines (coinciding with the start of the first wave of the COVID-19 pandemic in India, 85.4% of respondents (n = 379) felt that telemedicine use had increased; 11.7 % (n = 52) felt it had decreased; 2.3 % (n = 10) felt it remained the same; and 0.7 % (n = 3) were unsure. Of the respondents who felt the use of telemedicine increased, the majority felt that it increased by up to 50 % (Table 1).

**Table 1.** Perceived changes in the use of telemedicine after the release of the Telemedicine Practice Guidelines in India. Numbers in parentheses indicate the number of respondents.

| Perceived % increase or decrease from the start of the pandemic | Respondents (85.4 % [n = 379]) who felt telemedicine use increased in their: |  | Respondents (11.7 % [n = 52]) who felt telemedicine use decreased in their: |  |
| --- | --- | --- | --- | --- |
|  | *practice % (n)* | *region/hospital % (n)* | *practice % (n)* | *region/hospital % (n)* |
| < 25% | 50.4% (191) | 46.4% (176) | 21.2% (11) | 28.8% (15) |
| 25–50% | 34.6% (131) | 37.2% (141) | 44.2% (23) | 30.8% (16) |
| 50–75% | 12.4% (47) | 14.2% (54) | 23.8% (15) | 36.5% (19) |
| > 75% | 2.6% (10) | 2.1% (8) | 5.8% (3) | 3.8% (2) |
| **Total** | **100% (379)** | **100% (379)** | **100% (52)** | **100% (52)** |

Respondents were also asked about their views on whether telemedicine would and should play a significant part in their practice after the pandemic. Most respondents felt that telemedicine would continue to be a considerable part of their practice (78.4 % [n = 348]), while 13.5 % (n = 60) felt that it would not, and 8.1 % (n = 36) were unsure. Finally, 95.3 % of respondents believed telemedicine should play a substantial role in delivering healthcare in India in the future.

### 3.2 Challenges with telemedicine use in India after release of the Telemedicine Practice Guidelines

Survey responses (n = 115) to two optional open-ended questions on the challenges of telemedicine use in India (“What are the main challenges with Telemedicine that you have faced so far?”) and additional feedback were categorised into 11 codes agreed upon by the authors through an inductive process (Figure 1). The authors could not arrive at a consensus for one survey response, “Abuse of same prescription”.

Most responses (n = 51) were categorised under a code that indicated that telemedicine-led healthcare delivery compromised treatment quality. The second largest number of responses (n = 22) indicated that ‘Accessibility, quality and maturity of software and hardware infrastructure’ was an important challenge. Table 2 provides a sample of representative verbatim responses under these two codes.

**Table 2.** Representative responses for the two codes covering the greatest proportion of responses.

| **Code** | **Representative responses** |
| --- | --- |
| *Compromise to treatment quality* | "Being unable to physically/clinically examine patient to arrive at a comprehensive diagnosis."<br>"The inability to examine a patient is a big drawback as it can hinder our capability to diagnose a disease."<br>"Many times we need to physically examine the patient. Many problems are picked up when we palpate or auscultate the patients and many things are missed by the patients in their complaints which are vital for diagnosis."<br>"It's a blind opinion patient not examined"<br>"We play safe as we do not know the pts due to which best treatment not possible."<br>"Suboptimal clinical assessment!"<br>"Patient to doctor relationship is almost materialistic..."<br>"Change in patient-doctor relationship"<br>"Patient-doctor relationship gonna like customer-call centre relationship, it is a lazy practice."<br>"Misunderstandings and miscommunication can happen."<br>"Follow up feedback with the patients."<br>"Convincing people to get lab testing done." |
| *Accessibility, quality and maturity of software and hardware infrastructure* | "The technology penetration in my country is still limited , although people have smart phones , there usage is limited to surfing and social media"<br>"Net connectivity"<br>"Network issues"<br>"Also logistic issues in remote localities"<br>"Problems faced during video consultation"<br>"Some time the transfer of the image is of poor quality and that can affect the diagnosis" |
|  | "Sharing of past medical records" |

The remaining 42 responses were categorised under a variety of challenges, including limitation in the type of condition that can be managed (n = 8); governance issues (n = 7); inability or unwillingness of patients (n = 7 and 6, respectively) or doctors to use the system (n = 1 and 1, respectively); lack of awareness (n = 6); reimbursement issues (n = 4); and potential for excess healthcare utilisation (n = 2).

## 4. Discussion

Our survey was conducted in a period with a substantial push for nationwide use of telemedicine services. This was reflected in our survey, with a substantial portion of respondents stating that telemedicine usage had increased in their practice *and* region/hospital. This finding is also consistent with studies done globally (e.g., [24]) that compared healthcare workers’ opinions on telemedicine before and during the COVID-19 pandemic. The common theme across these studies was that despite the significant uptake, perceived benefits, and anticipated future role of telemedicine, numerous challenges must be addressed to realise telemedicine’s full potential. Our study provides a unique perspective on the nuances around these challenges, and they are also contextualised through the lens of the field experience of the study co-author (BGD), who actively engaged in the National Telemedicine Service (eSanjeevani) and was on the COVID-19 frontlines in India.

Reflecting on our survey responses and BGD’s experiences, three broad themes were identified regarding the challenges of telemedicine use in India: user experience-based integration, non-user-based integration challenges and technical challenges.

### 4.1 User experience-based integration challenges

Shifting to a completely non-physical consultation cannot be expected to be smooth because it contradicts how medicine has been practised for centuries—in-person, face-to-face, and with a clinician’s ‘healing’ touch. A stark manifestation of this is an unwillingness on the part of both patients and physicians to use telemedicine, though there were only seven responses that indicated this. Some responses also suggested an inability to use telemedicine, although training can overcome this challenge (see the section below on policy and practice).

A more significant set of challenges is linked to the doctor-patient relationship and the ability to do a physical examination, which were the most prevalent challenges identified in our survey. Besides having to unlearn how medicine is normally practised, it is important to recognise that physicians are more likely to uncover additional concerns during in-person consultations and examinations [25]. Furthermore, the lack of physical contact can also manifest as lost spaces in the consultation for patients to raise additional concerns (both clinical and non-clinical) [26]. These factors also negatively impact the ability to form and grow the doctor-patient relationship – as one of our respondents mentioned, “Patient-doctor relationship gonna like customer-call centre relationship, it is a lazy practice.”

### 4.2 Non-user-based integration challenges

A challenge that several respondents highlighted was that telemedicine was not appropriate for the management of all conditions/situations, and especially for emergencies – “Useful for mild health problems”. The authors feel, however, that this is more a *limitation* of telemedicine rather than a challenge, because it will never be appropriate for telemedicine to be used for these conditions/situations. Instead, it requires sound clinical judgement and better triage processes to ensure that telemedicine is *appropriately* used. Other non-user-based integration challenges are more significant.

For the use cases where telemedicine can be appropriate, its *inappropriate use* can be linked to defensive medicine, overdiagnosis, and overtreatment. One respondent implicated “playing safe” on the part of service providers in the absence of in-person physical examination and interaction as a key variable in compromising treatment quality. Another respondent referred to the problematic phenomenon of ‘doctor shopping’ – wherein patients fix multiple appointments with different doctors for the same complaint to get multiple opinions and/or prescriptions.

Seven respondents also felt that governance issues must be investigated, such as the legitimacy and inherent medicolegal risks of telemedicine practice, the legality and overuse of prescriptions, and the risk of unqualified/unlicensed or inadequately trained providers offering virtual services. Finally, it is essential to highlight that there is currently no provision in India for reimbursement from medical insurance for telemedicine practice [27].

### 4.3 Technical challenges

If users feel they cannot rely on telemedicine, they will not put their trust in the technology, and this will be reflected in less use. In our survey, 26% of responses were concerned about the lack of infrastructure to support telemedicine services, particularly in remote areas – “technology penetration in my country is still limited”, “logistic issues in remote localities”, and “network issues”. Other responses indicated the quality of image transmission and prohibitive hardware and software infrastructure costs as important factors. Respondents also highlighted the current limitations of telemedicine – namely, the ability to perform and interpret laboratory or imaging-led investigations and the lack of availability of past medical history and records.

## 5. Implications for policy and practice

Telemedicine implementation is challenging in low-resource settings for a variety of reasons, including: (a) infrastructural costs linked to set-up; (b) unreliable internet connectivity in rural areas; (c) slow clinical acceptance; (d) lack of conformance to standards; (e) non-interoperable systems; (f) non-user-friendly technologies; (g) lack of clearly articulated regulatory frameworks; and (h) faulty implementation strategies [28]. Despite these challenges (also) highlighted in our study, over 95% of our survey respondents felt telemedicine should play a substantial role in delivering healthcare in post-pandemic India. The authors suspect that a reason for this could be the recognition that telemedicine could serve as a ‘virtual bridge’ to universal and equitable healthcare access in India, where the majority of healthcare infrastructure is in urban centres despite most of the population living in rural areas [29]. Below, we highlight opportunities for integration of telemedicine in India using the same themes used to group the challenges. It is important to note that these opportunities are not mutually exclusive.

### 5.1 User experience-based integration opportunities

For telemedicine to become a standard component of care pathways, we will need to either see a change in norms of care delivery or telemedicine technology will have to evolve to better meet the requirements and expectations of patients and physicians – the reality will likely consist of both.

A starting point would be to explore telemedicine usage, adoption, and patient and clinician feedback (rural vs. urban-based) to better understand patient and clinician experiences and needs. There is, for example, evidence of the differential use of telemedicine among different groups according to various demographic attributes (e.g., rurality, region, literacy, etc.) [30,31]. Understanding care-seeking behaviour would be valuable in tailoring telemedicine services driven by a needs-led approach [23]. Further, it would be helpful to understand the shift in telemedicine adoption in different disciplines of medicine and healthcare and gauge their usage, efficacy, and impact.

Training and education of clinicians and patients will also help improve adoption and the ability of different users to easily adjust to care delivered through telemedicine [32. Furthermore, a hybrid telehealth model consisting of alternating in-person and telehealth visits could suit the Indian context well [33].

### 5.2 Non-user-based integration opportunities

One response that particularly resonated with the authors was that “there should be a national framework for referral” so that a patient is first seen by a general practitioner (GP) before referral to a specialist. General practitioners are significantly underrepresented in the current eSanjeevani referral pathway. This allows paramedics at the ‘spoke’ or individual patients to call specialist physicians at the state telemedicine ‘hubs’ directly – often for minor complaints, and they frequently use incomplete and vaguely typed messages to describe their complaints without any audio-visual input. The failure to account for these factors costs India’s healthcare system unnecessary resources, prevents other patients with more urgent needs from availing of appropriate appointment slots, and prevents the concerned specialist from providing informed and well-considered advice. As an example of this suboptimal use of resources, the study co-author, BGD, practices as an IVF specialist and has had several referrals coming to him of men with headaches. Implementing triage pathways, such as those used in the UK National Health Service (NHS), as well as linking every appointment to a patient’s ABHA/Aadhaar identifier while retaining options for second or even third opinions, where genuinely warranted, could help to optimise the use of telemedicine in India. Additionally, establishing an online ABHA/Aadhaar-tagged record of the disbursement of prescription drugs would help to curb the abuse of the same or multiple prescriptions for repeated access to habit-forming medications – a concern identified by one of the respondents.

Another critical factor highlighted in telemedicine integration is the need for further government involvement in funding and technical support, especially in rural settings, but this must be done with caution because of a lack of reliable data on quality as well as technical difficulties in assessing quality [34, 35]. There have also been several calls for specific legislation in India to tackle the various medicolegal issues associated with telemedicine and virtual care [27, 32].

Healthcare professionals and policymakers must consider these intersectional issues for the appropriateness of telemedicine implementation in the Indian context and also recognise the important disparities between India’s different states [36].

### 5.3 Technical opportunities

Scott Kruse et al. [37] highlight that the top barriers to telemedicine adoption are technology-specific (e.g., lack of computer literacy and technology barriers) that could be overcome through change-management techniques, training, and alternating care delivery by telemedicine and personal patient-to-provider interaction. Similar barriers influencing the implementation of telemedicine in India’s healthcare system (data privacy and security concerns, infrastructure issues) were outlined in a recent systematic review [38]. Further, Venkataraman et al. [38] emphasised the need for reliable internet connectivity, stringent data privacy measures, technological adoption, and positive sociocultural attitudes as vital facilitators for effective telemedicine adoption in India. India has made, and continues to make, substantial investments to improve its technological infrastructure, including a shift to 5G, as well as support for internet and smartphone growth in India in urban as well as rural areas. If designed, implemented and harnessed well, these initiatives could be essential to increasing and ensuring more appropriate use of telemedicine in India.

## 6. Study limitations

Several limitations decrease the study’s generalisability. The qualitative analysis of open-ended responses and their coding via inductive (and thematic analyses) approaches are typically associated with subjectivity and bias, which the authors (from diverse professional and academic backgrounds) have tried to minimise by following best practices and building consensus, as detailed in the Methods section.

## 7. Recommendations for future work

Surveying telemedicine usage, adoption, and patient feedback (rural versus urban and for patients with different conditions and presenting complaints) is warranted to understand the end-users, patient experience and the types of demand for services. This would be valuable in tailoring telemedicine services driven by a needs-led approach, as patient experience is often measured using tools specific to the clinical population or context [23]. It would be helpful to understand the shift in telemedicine adoption in different disciplines of medicine and healthcare and gauge their efficacy and impact. Finally, focusing on addressing the key perceived challenges of ‘compromise to treatment quality’ and ‘accessibility, quality and maturity of software and hardware infrastructure’ needs to be investigated. For the former, a more robust triaging process can ensure that only those patients who can be helped by telemedicine-based care are presented with this option. For the latter, while India has made great strides in improving its digital infrastructure, there is still much that needs to be done to ensure the ‘last mile’ and equitable delivery of care [39–41]. Recently, Wiley et al. [42] have concluded that organisational, environmental and personal facilitators drive quality perceptions among physicians. This will obviously require greater resources, but before that, it requires that considerations and tough decisions are made on trade-offs and opportunity costs linked with investment in digital infrastructure at the expense of other areas that may have a greater impact on population health, like education, housing, access to clean water and access to good nutrition. On a related note, standardised cost-effectiveness studies comparing telehealth with in-person care should be conducted to strengthen the evidence base before extensive implementation across various setting [41, 43]. Finally, Ummer et al. (2025) have noted the need to improve the evidence base on telemedicine’s impact in India across key aspects such as quality of care, care-seeking behaviour, cost implications and changes in health outcomes. nutrition.

## 8. Conclusions

Despite the considerable uptake, perceived benefits, and the foreseen positive role of telemedicine in India, several challenges of telemedicine use (viz., technical, user experience-based integration, and non-user-based integration) have been identified through our study, which need to be addressed to realise telemedicine’s potential. Several relevant opportunities are suggested to inform policy and practice to ensure effective utilisation of telemedicine in India.

As healthcare in India undergoes unprecedented transformation, our study provides insights to support effective telemedicine adoption and integration into existing care pathways. Supported by favourable and opportune policies (especially the Telemedicine Practice Guidelines [11]) by the Indian government, telemedicine played a crucial role in offering healthcare access during the COVID-19 pandemic. Overcoming the structural issues of the Indian healthcare system can position it to play a transformative role in ensuring universal access to healthcare in India [18,29]. Our findings also contribute to the growing body of work to support stakeholders across the healthcare sector in India (and other low-resource settings) to understand factors impacting the implementation and optimisation of telemedicine utilisation in an ever-evolving healthcare and technological landscape.

## Data Availability

All data produced in the present study are available upon reasonable request to the authors

## Acknowledgements

The authors thank the Oxford India Centre for Sustainable Development team, particularly Ms Vinita Govindarajan (Partnerships & Communications Manager), for their help with survey dissemination and valuable advice. Additionally, the authors would like to acknowledge Dr Santanu Sen (former President of the Indian Medical Association), Dr Surekha Patil (Dean, D.Y. Patil University School of Medicine), Dr Sriram Gopal (Head of Ob-Gyn Department, D.Y. Patil University School of Medicine), and Dr Vivekananda Bheemisetty (Orthopaedic surgeon at Zoi Hospitals, Hyderabad) for support with survey dissemination across India. Finally, the authors would like to express their profound gratitude to Dr Vijay Patil (President of D.Y. Patil University) for supporting this study.

## Author contributions

ARJ and VHN conceived the project; VHN and ARJ conceptualised the methodology with contributions from BGD; VHN designed and implemented the internet-based questionnaire with contributions from ARJ and BGD; BGD, VHN, and ARJ undertook survey dissemination; VHN collected and curated the data; VHN, BGD, SS, and ARJ contributed to formal analysis, investigation, visualisation, writing of final results, and preparation of the original draft manuscript; VHN and ARJ undertook supervision, project administration, and ensuring requisite infrastructure; all authors contributed to the subsequent draft review and editing; all authors have read and agreed to the final version of the manuscript.

## Ethics approval

This is a service review. The Oxford Tropical Research Ethics Committee has confirmed that no ethical approval is required for this research.

## Declarations

None

## Declaration of generative AI in scientific writing

None

## Declarations

None

## References

1. Sieck CJ, Sheon A, Ancker JS, et al. Digital Inclusion as a social determinant of health. npj Digit. Med. 2021; 4: 52.

2. Kickbusch I, Piselli D, Agrawal A, et al. The Lancet and Financial Times Commission on governing health futures 2030: Growing up in a Digital World. Lancet 2021; 398: 1727–1776.

3. Richardson S, Lawrence K, Schoenthaler AM, et al. A framework for Digital Health Equity. npj Digit. Med. 2022; 5(1), 119.

4. World Health Organization. What you need to know about Digital Health Systems. World Health Organization. https://www.who.int/news/item/05-02-2019-what-you-need-to-know-about-digital-health-systems (2019, accessed 21 December 2023).

5. McKinsey & Company. Digital India - McKinsey & Company. Digital India: Technology to transform a connected nation. https://www.mckinsey.com/∼/media/mckinsey/business%20functions/mckinsey%20digital/our%20insights/digital%20india%20technology%20to%20transform%20a%20connected%20nation/digital-india-technology-to-transform-a-connected-nation-full-report.pdf (2019, accessed 21 December 2023).

6. Ministry of Electronics & Information Technology. Digital India. https://digitalindia.gov.in/ (2023, accessed 21 December 2023).

7. Ernst & Young India. Digitalizing India: A force to reckon with. EY US – Home. https://www.ey.com/en_in/india-at-100/digitalizing-india-a-force-to-reckon-with (2023, accessed 21 December 2023).

8. Deloitte. Technology, Media, and Telecommunications (TMT) predictions 2022. Deloitte TMT Predictions 2022. https://www2.deloitte.com/content/dam/Deloitte/in/Documents/technology-media-telecommunications/in-TMT-predictions-2022-noexp.pdf (2022, accessed 21 December 2023).

9. National Digital Health Mission. Ministry of Health and Family Welfare, Government of India. National Digital Health Mission – NITI Aayog. Strategy Overview – Making India a Digital Health Nation Enabling Digital Healthcare for all. https://www.niti.gov.in/sites/default/files/2023-02/ndhm_strategy_overview.pdf (2020, accessed 21 December 2023).

10. National Digital Health Blueprint – ABDM. Ministry of Health and Family Welfare, Government of India. National Digital Health Blueprint. https://abdm.gov.in:8081/uploads/ndhb_1_56ec695bc8.pdf (2017, accessed 21 December 2023).

11. MOHFW. Telemedicine practice guidelines – MOHFW. Telemedicine Practice Guidelines - Enabling Registered Medical Practitioners to Provide Healthcare Using Telemedicine. https://www.mohfw.gov.in/pdf/Telemedicine.pdf (2020, accessed 21 December 2023).

12. Ministry of Electronics & Information Technology. Esanjeevani. India Global Stack. https://www.indiastack.global/esanjeevani/ (2020, accessed 21 December 2023).

13. Kaboré SS, Ngangue P, Soubeiga D, et al. Barriers and facilitators for the sustainability of digital health interventions in low and middle-income countries: A systematic review. Front. Digit. Health. 2022; 4: 1014375.

14. Cross M. Doctors are reluctant to use telemedicine and misunderstand what patients want, says NHS Confederation Report. BMJ 2011; 342.

15. Justinia T. The UK’s National Programme for it: Why was it dismantled? Health Serv Manage Res. 2017; 30: 2–9.

16. Eysenbach G. Improving the quality of web surveys: The checklist for reporting results of internet E-surveys (CHERRIES). J. Med. Internet Res. 2004; 6: 132.

17. Kelley K, Clark B, Brown V, Sitzia J. Good practice in the conduct and reporting of survey research. Int J Qual Health Care 2003; 15: 261–6.

18. Nagaraja VH, Ghosh Dastidar B, Suri S, Jani A. Perspectives and use of telemedicine by doctors in India: A cross-sectional study. Health Policy Technol. 2024; 100845.

19. Pope C, Ziebland S, and Mays N. Analysing qualitative data. BMJ 2000; 320: 114–116.

20. Braun V and Clarke V. Using thematic analysis in psychology. Qual Res Psychol. 2006; 3: 77–101.

21. Nowell LS, Norris JM, White DE, Moules NJ. Thematic analysis: Striving to meet the trustworthiness criteria. Int. J. Qual. Methods 2017; 16: 1609406917733847.

22. Vaismoradi M, Turunen H, Bondas T. Content analysis and thematic analysis: Implications for conducting a qualitative descriptive study. Nurs Health Sci 2013; 15: 398–405.

23. Langbecker D, Caffery LJ, Gillespie N and Smith AC. Using survey methods in telehealth research: A practical guide. J Telemed Telecare 2017; 23: 770–779.

24. Nitiema P. Telehealth before and during the COVID-19 pandemic: analysis of health care workers’ opinions. J. Med. Internet Res. 2022; 24: e29519.

25. White AEC. When and how do surgeons initiate noticings of additional concerns?. Soc. Sci. Med. 2020; 244: 112320.

26. White SJ, Nguyen A, Cartmill JA. Agency and the telephone: patient contributions to the clinical and interactional agendas in telehealth consultations. Patient Educ Couns 2022; 105: 2074–2080.

27. Ateriya N, Saraf A, Meshram VP, Setia P. Telemedicine and virtual consultation: The Indian perspective. Natl Med J India 2018; 31.

28. Combi C, Pozzani G, Pozzi G. Telemedicine for developing countries. A survey and some design issues. Appl Clin Inform 2016; 7: 1025.

29. Ghosh Dastidar B, Suri S, Nagaraja VH, Jani A. A virtual bridge to Universal Healthcare in India. Commun. Med. 2022; 2: 145.

30. Pierce RP and Stevermer JJ. Disparities in the use of telehealth at the onset of the COVID-19 public health emergency. J Telemed Telecare 2023; 29: 3–9.

31. Chu C, Cram P, Pang A, Stamenova V, Tadrous M, Bhatia RS. Rural telemedicine use before and during the COVID-19 pandemic: repeated cross-sectional study. J. Med. Internet Res. 2021; 23: e26960.

32. Datta R, Singh A, Mishra P. A survey of awareness, knowledge, attitude, and skills of telemedicine among healthcare professionals in India. Med J Armed Forces India. 2023;79(6), pp.702–709.

33. Raj Westwood A. Is hybrid telehealth model the next step for private healthcare in India?. Health Serv. Insights 2021; 11786329211043301.

34. Jarosławski S. and Saberwal G. In eHealth in India today, the nature of work, the challenges and the finances: an interview-based study. BMC Med. Inform. Decis. Mak. 2014; 14: 1–12.

35. Mohanan M. Hay K, Mor N. Quality of health care in India: challenges, priorities, and the road ahead. Health Aff. 2016; 35: 1753–1758.

36. Dhyani VS, Krishnan JB, Mathias EG, et al. Barriers and facilitators for the adoption of telemedicine services in low-income and middle-income countries: a rapid overview of reviews. BMJ Innov. 2023; 9: 215–225.

37. Scott Kruse C, Karem P, Shifflett K, Vegi L, Ravi K, Brooks M. Evaluating barriers to adopting telemedicine worldwide: a systematic review. J Telemed Telecare. 2018; 24: 4–12.

38. Venkataraman A, Fatma N, Edirippulige S, Ramamohan V. Facilitators and Barriers for Telemedicine Systems in India from Multiple Stakeholder Perspectives and Settings: A Systematic Review. Telemed E-Health. 2024. DOI: 10.1089/tmj.2023.0297

39. Sood S, Lal K, Bhatia M, Kapoor G, Singh S, Kaushish RK, Bisht D, Sharma VK, Chakraborty S, Shete M, Solanki NK. Adoption & Utilization of India’s eSanjeevani National Telemedicine Service. Oxf. Open Digit. Health. 2025; p.oqaf025

40. Hussain N. Digital Health Inequities: Analysing Barriers to Access Across Demographic Groups in India. JDPP. 2025. p.24551333251382726

41. Parthasarathi A, George T, Kalimuth MB, Jayasimha S, Ullah MK, Patil R, Nair A, Pai U, Inbarani E, Jacob AG, Chandy VJ.Exploring the potential of telemedicine for improved primary healthcare in India: a comprehensive review. Lancet Reg. Health - Southeast Asia. 2024;27.

42. Wiley K, Pugh A, Brown-Podgorski BL, Jackson JR, McSwain D. Associations between telemedicine use barriers, organizational factors, and physician perceptions of care quality. Telemed e-Health. 2024;30(12), pp.2883–2889.

43. Shambushankar AK, Jose J, Gnanasekaran S, Kaur G, KS, A. Cost-Effectiveness of Telerehabilitation Compared to Traditional In-Person Rehabilitation: A Systematic Review and Meta-Analysis. Cureus. 2025; 17(2).

44. Ummer O, Sarangi A, Khanna A, Mohan D, Scott K, LeFevre A. Lessons Learned From Over 20 Years of Telemedicine Services in India: Scoping Review of Telemedicine Services Initiated From 2000 to 2023. J. Med. Internet Res. 2025; 27, p.e63984

